# Improving COVID-19 critical care mortality over time in England: A national cohort study, March to June 2020

**DOI:** 10.1101/2020.07.30.20165134

**Authors:** John M. Dennis, Andrew P. McGovern, Sebastian J. Vollmer, Bilal A. Mateen

**Author notes:** **Author for Correspondence** Dr. Bilal A. Mateen, Social Science and Systems in Health Unit, Warwick Medical School, University of Warwick, Coventry, CV4 7AL, UK.

## Abstract

**Objectives:** To determine the trend in mortality risk over time in people with severe COVID-19 requiring critical care (high intensive unit [HDU] or intensive care unit [ICU]) management.

**Methods:** We accessed national English data on all adult COVID-19 specific critical care admissions from the COVID-19 Hospitalisation in England Surveillance System (CHESS), up to the 29th June 2020 (n=14,958). The study period was 1st March until 30th May, meaning every patient had 30 days of potential follow-up available. The primary outcome was in-hospital 30-day all-cause mortality. Hazard ratios for mortality were estimated for those admitted each week using a Cox proportional hazards models, adjusting for age (non-linear restricted cubic spline), sex, ethnicity, comorbidities, and geographical region.

**Results:** 30-day mortality peaked for people admitted to critical care in early April (peak 29.1% for HDU, 41.5% for ICU). There was subsequently a sustained decrease in mortality risk until the end of the study period. As a linear trend from the first week of April, adjusted mortality risk decreased by 11.2% (adjusted HR 0.89 [95% CI 0.87 - 0.91]) per week in HDU, and 9.0% (adjusted HR 0.91 [95% CI 0.88 - 0.94]) in ICU.

**Conclusions:** There has been a substantial mortality improvement in people admitted to critical care with COVID-19 in England, with markedly lower mortality in people admitted in mid-April and May compared to earlier in the pandemic. This trend remains after adjustment for patient demographics and comorbidities suggesting this improvement is not due to changing patient characteristics. Possible causes include the introduction of effective treatments as part of clinical trials and a falling critical care burden.

## Introduction

Recent national data from England suggests there has been a substantial reduction in coronavirus disease 2019 (COVID-19) in-hospital mortality over recent months, with an estimated decline in the case-fatality rate of hospitalised patients from 6% in early April to 1.5% by mid-June.[1] One potential explanation for this observation is a shift in the demographics of people admitted with COVID-19 towards younger and overall less comorbid individuals, resulting in a lower risk of mortality.[2] Another possibility is that expanded testing has led to increased identification of milder cases. If the latter is a significant factor in the declining mortality then we would expect mortality in the critical care setting to have remained relatively unchanged.

We sought to test whether the same trend of improving mortality over time has been seen in people with severe COVID-19 requiring critical care (high intensive unit [HDU] or intensive care unit [ICU]) management, whether falling mortality reflected changes in patient demographics, and whether time trends varied by demographics and geography in England.

## Methods

### Data Source

We accessed national English data from the COVID-19 Hospitalisation in England Surveillance System (CHESS),[3] a statutory collection of all individuals with confirmed or clinically presumed COVID-19 managed in HDU or ICU. CHESS data were extracted on 29th June, 2020. Our study period was 1st March until 30th May, meaning every patient had 30 days of potential follow-up available. People aged 18-99 were eligible, pregnant women (n=88) were excluded. Two cohorts were defined; 1: all people admitted to HDU but never ICU; 2: all people admitted to ICU.

### Recorded clinical features

CHESS contains individual-level demographic characteristics: age, sex, ethnicity, admitting hospital, and recorded comorbidities (diabetes; asthma; other chronic respiratory disease; chronic heart disease; hypertension; immunosuppression due to disease or treatment; chronic neurological disease; chronic renal disease; and chronic liver disease). We coded ethnicity as white, Asian, black, mixed, and other, and categorised hospital centres by region: London, East of England, Midlands, North East and Yorkshire, North West, South East, and South West.

### Statistical Analysis

The primary outcome was in-hospital 30-day all-cause mortality. To account for discharge from hospital as a competing risk, people who were discharged or transferred were assumed no longer at risk of death and so were not censored at the day of discharge and instead had their follow-up time set to 30 days.[4] We estimated adjusted hazard ratios for mortality by week of admission using adjusted Cox proportional hazards models, adjusting for age (non-linear restricted cubic spline), sex, ethnicity, comorbidities, and geographical region, with proportional hazards assumptions tested. For the ICU cohort, follow-up began at the date of ICU admission with additional adjustment for the number of days from hospital to ICU admission. After initial analysis modelling week of admission as a categorical variable, we ran additional models with week of admission from the week of 29th March, 2020 to 24th May, 2020 modelled as a linear term. As sensitivity analysis we repeated models with National Health Service (NHS) hospital trust included as a random effect.

## Results

14,958 individuals (HDU n=10,479; ICU n=4,479) were eligible for the study, of whom 4,474 (29.9%) died (HDU n=2,733 [24.2%]; ICU n=1,741 [36.1%]). CHESS demographics have been previously reported,[5] and are therefore not replicated here. Peak unadjusted mortality was 29.1% for HDU (people admitted in week 4) and 41.4% for ICU (week 5). Lowest mortality was seen towards the end of follow-up; 10.2% for HDU (week 13) and 24.8% for ICU (week 11).

Adjusted mortality increased for people admitted to both HDU and ICU from 1st March, 2020, up to the first week of April [Figure 1]. Subsequently, there was a sustained decrease in mortality risk until the end of the study period. These trends were not substantially modified following adjustment for NHS hospital trust as a random effect. As a linear trend from the first week of April, adjusted mortality risk decreased by 11.2% (adjusted HR 0.89 [95% CI 0.87 - 0.91]) per week in HDU, and 9.0% (adjusted HR 0.91 [95% CI 0.88 - 0.94]) in ICU.

**Figure A:**
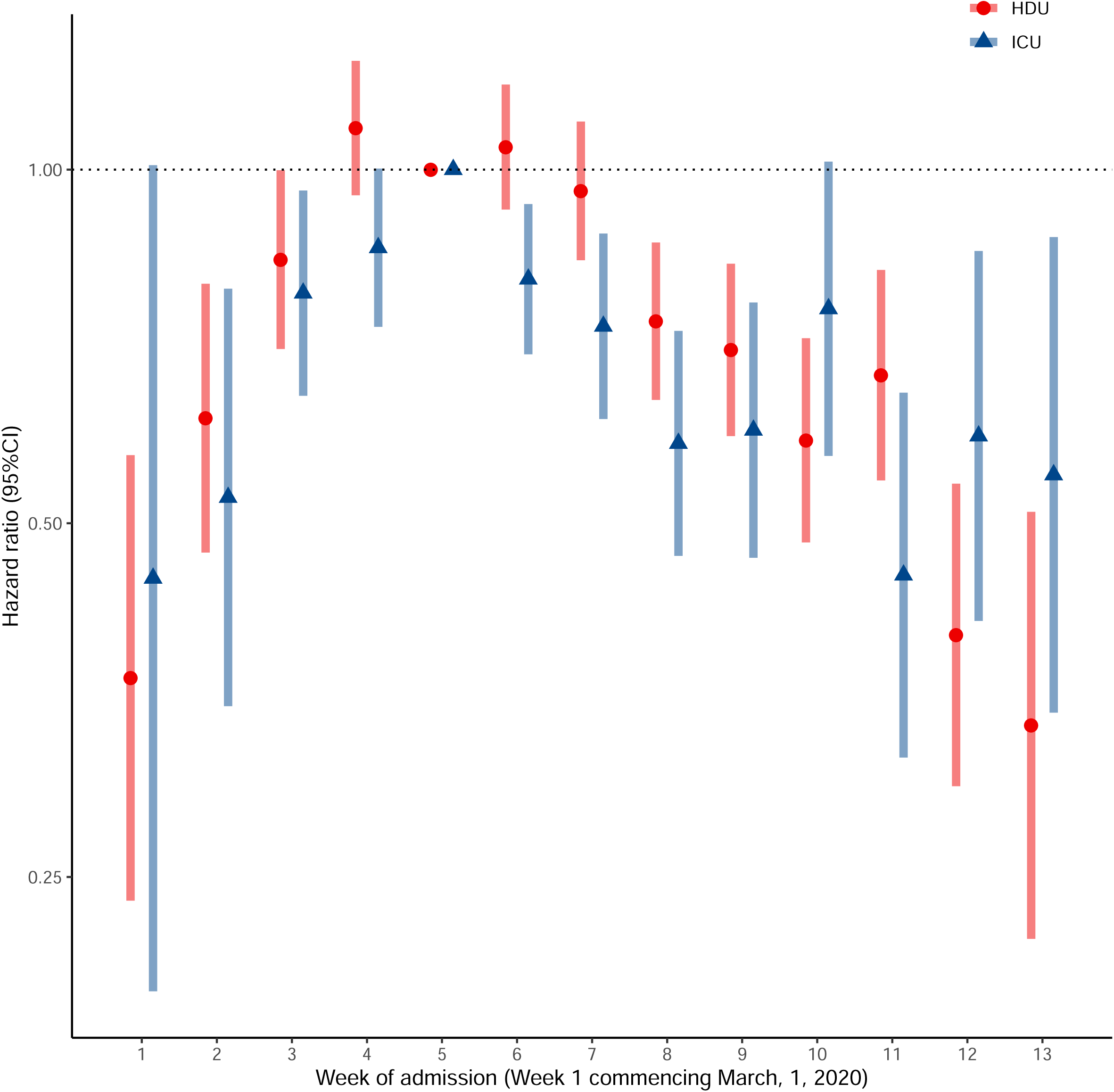
Adjusted hazard ratios for the weekly time trend for in-hospital mortality for people admitted to critical care with COVID-19 in England, from the week of 1st March, 2020 to the week of 24th May, 2020. Results are shown separately for people admitted to HDU and ICU. A hazard ratio less than 1 reflects a lower mortality compared to week 5 (the peak week of admissions in CHESS). Week 5 (commencing 29th March, 2020) was used as the reference group as it was the peak week of COVID-19 admissions in CHESS. Bars represent 95% confidence intervals.

**Figure 1B:**
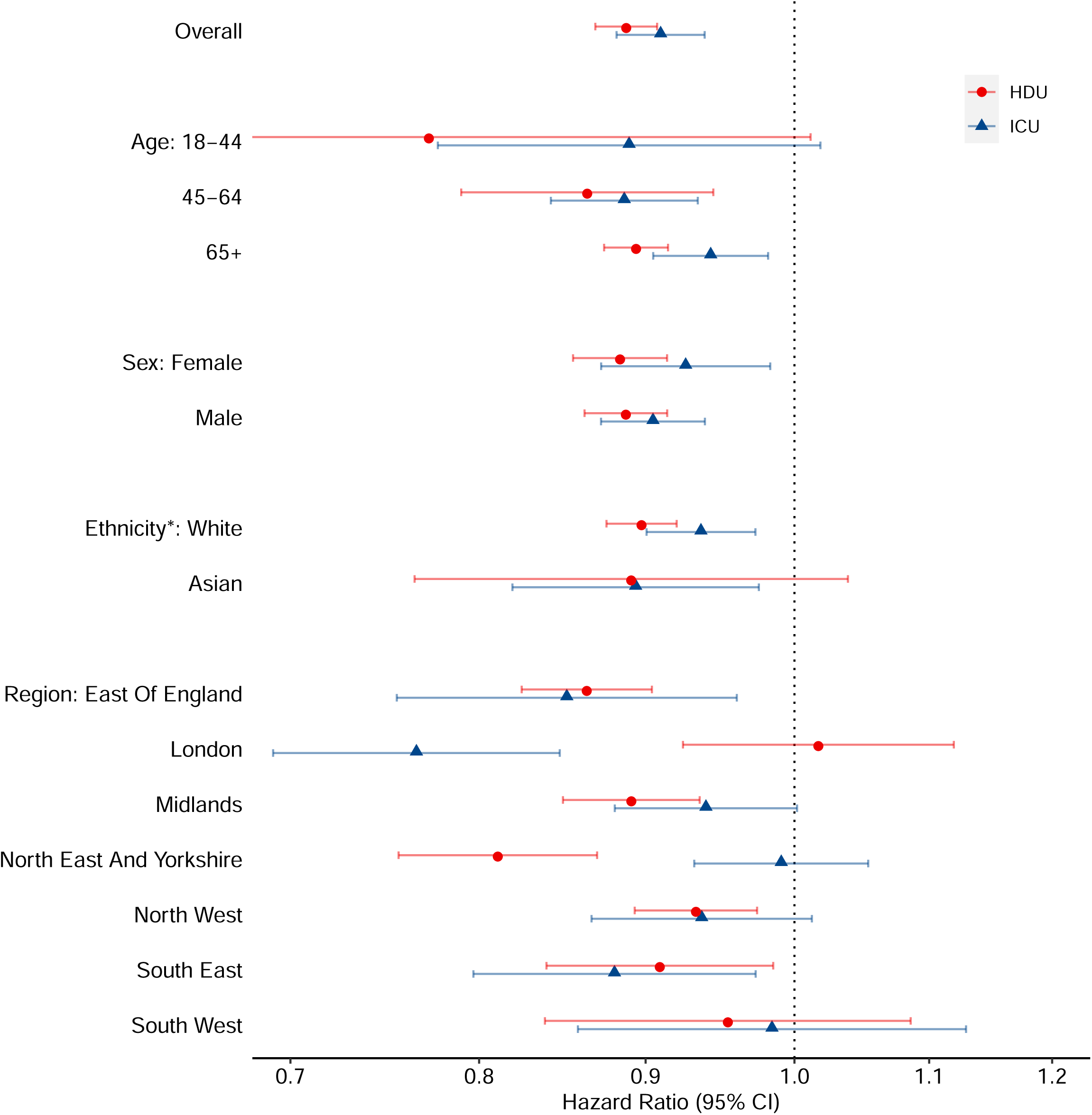
Adjusted hazard ratios for the linear time trend for in-hospital mortality for people admitted to critical care with COVID-19 in England, from 29th March to 29th May, 2020. Results are shown for HDU and ICU admissions, overall and by subgroup. A hazard ratio less than 1 reflects a mortality improvement over the study period. Bars represent 95% confidence intervals. *Hazard ratios for other ethnicity defined subgroups not plotted due to low numbers: Black (HDU HR 0.53 [95% CI: 0.36 - 0.81], ICU HR 0.92 [95% CI: 0.78 - 1.08]); Mixed (HDU HR 0.72 [95% CI: 0.49 - 1.07], ICU HR 0.72 [95% CI: 0.55 - 0.95]); Other (HDU HR 0.97 [95% CI: 0.62 - 1.55], ICU HR 0.82 [95% CI: 0.68 - 0.99])

The linear time trend in mortality improvement from the week of 29th March, 2020 is consistent across subgroups defined by age, sex and ethnicity (Figure 1B). There is evidence of a strong regional effect, with London hospitals reporting a marked reduction in ICU mortality but no improvement in HDU mortality, and the opposite pattern seen in hospitals in the North East and Yorkshire (Figure 1B). Mortality improvements were observed in patient subgroups defined by the presence of diabetes (HDU HR 0.84 [95% CI: 0.79 - 0.89]; ICU HR 0.94 [95% CI: 0.89 - 1.00]) and chronic renal disease (HDU HR 0.92 [0.87 - 0.98]; ICU HR 0.87 [95% CI: 0.78 - 0.97]), but there was no evidence of an improvement for people with chronic respiratory disease admitted to ICU (HDU HR 0.91 [95% CI: 0.86 - 0.97]; ICU HR 0.98 [95% CI: 0.90 - 1.08]).

## Discussion

Our analysis, using the largest available COVID-19 specific national critical care database, shows a substantial recent improvement in mortality for people admitted to critical care with COVID-19 in England, with markedly lower mortality in people admitted in mid-April and May compared to earlier in the pandemic. Adjustment for all recorded patient level demographic and clinical features suggests this improvement does not reflect a change in patient demographics or comorbidities.

We observed an improvement in mortality for all regions in England, although interesting differences between changes in HDU and ICU specific mortality were seen in London and in the North East. A recent analysis of the CHESS dataset matched with national level audit data demonstrated that this marked variation in mortality by hospital trust was not due to a time-varying trend in the severity profile (i.e. APACHE-II score) of those admitted to ITU for COVID-19 related illnesses.[6] The combination of these two results suggest that further study is warranted to understand differences in mortality by hospital and geographical region across England.

A potential limitation of this analysis is the possibility of reporting delays leading to incomplete ascertainment of death in more recent weeks, although this was mitigated by our study design which focused on in-hospital mortality and ensured all eligible patients had at least 30 days follow-up available. Whilst reporting delays might plausibly lead to under-ascertainment of mortality for people admitted in May, a clear mortality improvement was also observed in April. As CHESS is a daily mandatory collection for all hospitals in England we would expect reasonably accurate and timely capture of in-hospital mortality, and so feel it is unlikely our findings simply reflect reporting delays.

Temporal changes in COVID-19 disease severity at admission, patient selection for critical care management, critical care treatment, hospital capacity, and COVID-19 testing, all offer potential explanations for our findings. Of particular note regarding treatment, the RECOVERY trial began nationwide recruitment in early April, and included interventions later shown to have mortality-specific, or length of ITU admission-specific benefits;[7] it is plausible recruitment to the trial might partially explain improved patient outcomes. Regarding capacity, it has been shown that bed saturation across England was at its highest in early April, and then progressively improved over the course of the 1st wave of the pandemic.[8] Therefore, the observed time trend could be a manifestation of the well established decline in patient-specific outcomes often observed at ‘unsafe’ occupancy levels.[9] Further interrogation of possible quality-of-care based explanations is required.

In conclusion, there has been a substantial mortality improvement in people admitted to critical care with COVID-19 in England, with markedly lower mortality in people admitted in mid-April and May compared to earlier in the pandemic. This trend remains after adjustment for patient demographics and comorbidities suggesting this improvement is not due to changing patient characteristics. Possible causes include the introduction of effective treatments as part of the RECOVERY trial and a falling critical care burden.

## Data Availability

Data cannot be shared publicly as it was collected by Public Health England (PHE) as part of their statutory responsibilities, which allows them to process patient confidential data without explicit patient consent. Data utilised in this study were made available through an agreement between the University of Warwick and PHE. Individual requests for access to CHESS data are considered directly by PHE (contact via)

## Acknowledgements

JMD is supported by an Independent Fellowship funded by Research England’s Expanding Excellence in England (E3) fund. SJV, and BAM are supported by The Alan Turing Institute (EPSRC grant EP/N510129/). SJV is supported by the University of Warwick IAA funding.

## Contributors

JMD and BAM designed the study. JMD wrote the analysis script on dummy code, with input from SJV and BAM. SJV ran the analysis. BAM, APM and JMD drafted the article. All authors provided support for the analysis and interpretation of results, critically revised the article, and approved the final article.

## Ethics & Governance

The study was reviewed and approved by the Warwick BSREC (BSREC 119/19-20-V1.1) and sponsorship is being provided by University of Warwick (SOC.28/19-20).

## Guarantor Statement

The senior authors (BAM and SJV) had full access to all data and had final responsibility for the decision to submit for publication.

## Patient and Public Involvement

No patients were involved in the design, interpretation of the results, or dissemination of this study.

## Patient consent for publication

Not required.

